# Smoking increases the risk of COVID-19 positivity, while Never-smoking reduces the risk

**DOI:** 10.1101/2020.11.30.20241380

**Authors:** Samson Barasa, Josephine Kiage-Mokaya, Katya Cruz-Madrid, Michael Friedlander

## Abstract

**Introduction:** Does smoking decrease the risk of testing positive for COVID-19 because the never-smokers (84%) prevalence is high and the current-smokers prevalence is low among COVID-19 positive patients? ^1,2,3,4,5,6^ We sought to determine whether never smoking increases the risk of COVID-19 positivity among the 50 to 69-year old patients because they are more likely to test positive for COVID-19.^7^

**Method:** We conducted a retrospective chart review of COVID-19 Polymerase chain reaction, in-hospital tested ≥18-year-old patients. A Poisson regression analysis stratified into never-smokers and history of smoking (current + former smokers) was conducted.

**Results:** 277 COVID-19 negative and 117 COVID-19 positive patients’ charts with a never-smokers prevalence of 42.32% and 54% respectively were analyzed. The never-smokers prevalence was 54%, 20-39-years; 61 %, 40 -49-years; 41%, 50 – 69-years; and 43%, 70 – 100-years.

The 40-49-year-old current and former smokers were more likely to test positive for COVID-19 [1.309 (1.047 - 1.635)], unlike the 40-49-year-old never-smokers [0.976 (0.890-1.071)] who had a lower risk.

Regardless of their smoking status, males [1.084(1.021 - 1.151)] and the 50-69-year-old patients [1.082 (1.014 -1.154)] were more likely to test positive for COVID-19, while end stage renal disease [0.908(0.843-0.978)] and non-COVID-19 respiratory viral illness [0.907 (0.863 - 0.953)] patients had a lower risk of COVID-19 positivity.

Heart failure [0.907 (0.830 - 0.991)], chronic obstructive pulmonary disease (COPD) [0.842 (0.745 - 0.952)] and Parkinson’s disease [0.823 (0.708 - 0.957)] never-smokers were less likely to test positive for COVID-19.

**Conclusion:** This is the first study to show that smoking increases the risk of COVID-19 positivity among the 40-49-year-old patients, while not smoking reduces the risk of COVID-19 positivity among the heart failure, COPD and Parkinson’s disease patients. This study emphasizes that COVID-19 positivity risk is not reduced by smoking and not increased by not smoking.

## Introduction

The association between smoking and COVID-19 infection is unclear. It has been speculated that smoking reduces the risk of testing positive for COVID-19 because the prevalence of current and former smokers is low among COVID-19 positive patients, while the prevalence of never smokers is high among COVID-19 positive patients. ^1,2, 3,4,5,6^

On the other hand, an increased risk of morbidity, mortality and need for mechanical ventilator support has been reported among COVID-19 positive current and former smokers. ^2^

The 50-69-year-olds are more likely to test positive for COVID-19.^7^ If smoking reduces the risk of testing positive for COVID-19, is the prevalence of current-smokers low and or the prevalence of never-smokers high among the 50-69-year-olds?

Does never smoking really increase or decrease the risk of testing positive for COVID-19 among hospitalized patients?

We sought to determine whether smoking increases the risk of testing positive for COVID-19 among the 50-69-year-old patients by stratifying patients into never smokers and those with history of smoking (current and former smokers).

## Methodology

### Study population and setting

A retrospective chart review of patients who were tested for COVID-19 during hospitalization using the PCR test, between 2^nd^ February 2020 and 21^st^ October 2020 was conducted in Eugene, Oregon United States of America (USA). The study enrollees were 18 years and older. The PeaceHealth institutional review board (IRB) approved the study and waived the written informed consent requirement.

### Data collection

The Microsoft Access software was used to manage data after being extracted from the electronic medical records.

### Risk factors

Data was obtained from documents and laboratory tests completed during hospitalization. Lymphopenia data was gathered from the complete blood count results. While the non-COVID-19 respiratory viral illness data was collected from the respiratory viral panel results. In addition, the following risk factors were examined: age, diabetes mellitus, stroke, dementia, congestive heart failure (CHF), COPD, asthma, systemic lupus erythromatosus (SLE), organ transplant, chronic kidney disease stage 3 (CKD-3), end stage renal disease (ESRD), coronary artery disease (CAD), smoking, hypertension, Parkinson’s disease, multiple sclerosis, demyelination disease, human immune-deficiency virus (HIV) infection and body mass index (BMI.

### Outcome

The study outcome was the COVID-19 PCR test result.

### Statistical analysis

Our study hypothesized that the 50-69-year old current plus former-smokers had a higher risk of testing positive for COVID-19 than the 50-to 69-year-old never-smokers.

Data was stratified into never-smokers versus history of smoking (current plus former smokers) and analyzed using the Generalized Poisson regression analysis while adjusting for the aforementioned risk factors. The statistical analysis was performed using STATA version 15.

## Results

### Demographics

The median and interquartile age for COVID-19 positive and negative patients was not significantly different. Similarly, gender distribution was not significantly different between COVID-19 positive and negative patients.

### Medical comorbidities

The prevalence of medical comorbidities was not significantly different between COVID-19 positive and negative patients, apart from COVID-19 positive patients having a significantly lower prevalence of ESRD, COPD, CHF and non-COVID-19 respiratory viral illness.

Both COVID-19 positive and negative patients had a high prevalence of lymphopenia, although the lymphopenia among the COVID-19 positive patients was significantly higher.

### Smoking and COVID-19

Figure 1 below shows that while COVID-19 positive patients had a significantly lower prevalence of current smokers (9%), they also had a higher prevalence of never smokers (54%) compared to COVID-19 negative patients. Both COVID-19 positive (37 %) and negative (39.33%) patients had an almost equal former-smokers’ prevalence.

**Figure 1.**
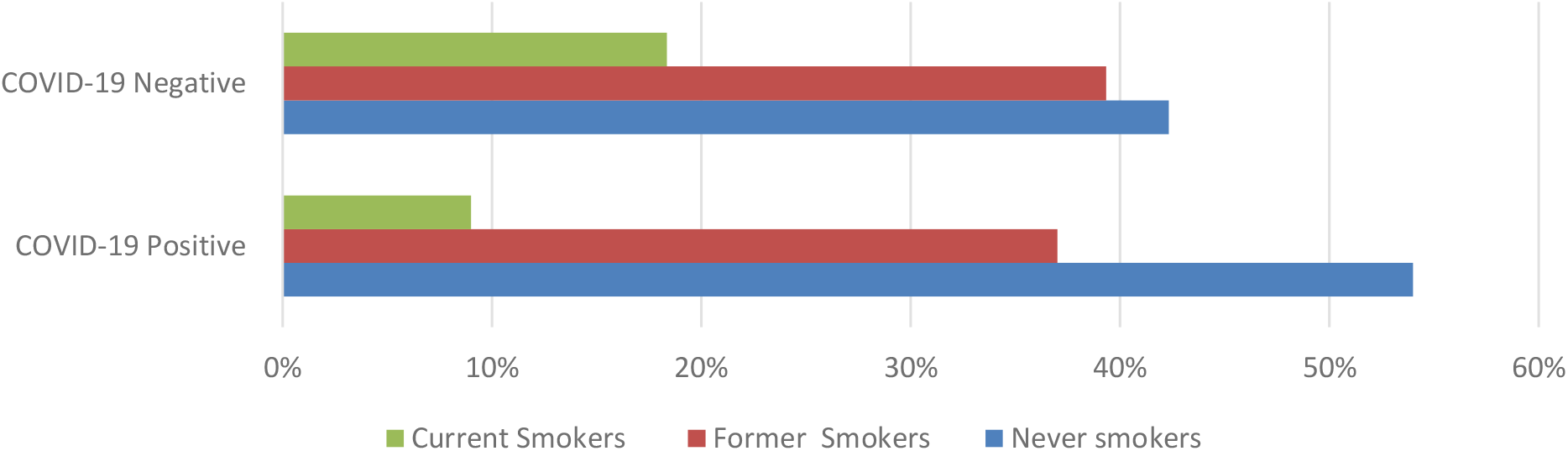
A graph of Never, Former and Current Smokers among COVID-19 Positive and Negative patients

Figure 2 below shows that the 40-49-year-old patients had the highest (60.98%) and lowest (14.63%) prevalence of never and former smokers respectively. In contrast, the 70-100-year-old patients had the lowest (4.79%) and highest (52.05%) prevalence of current and former smokers respectively. The prevalence of current smokers was almost equal among the 20-39-year-old (22%), 40-49-year-old (24%) and 50-69-year-old (23%) patients.

**Figure 2.**
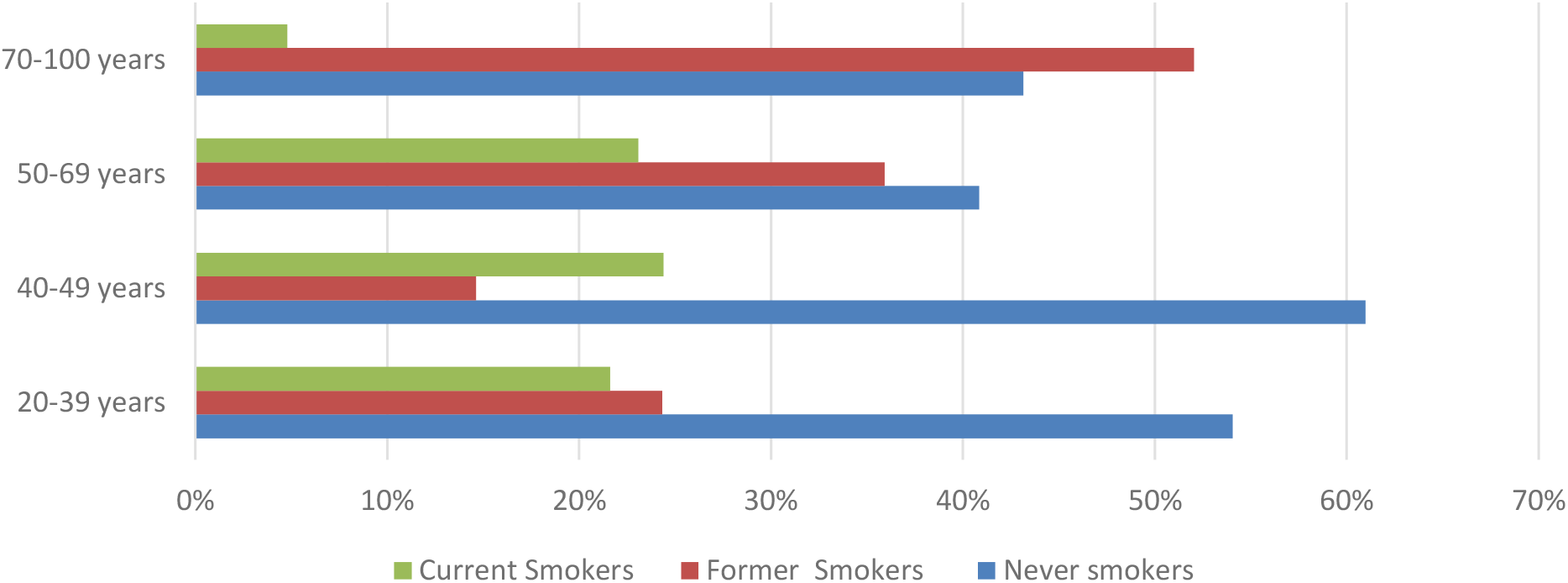
The graph of Never, Former and Current smokers across different Age groups

#### Analysis of COVID-19 status and age controlling for never smoking

Section1 of table 2 below shows partial results of COVID-19 status and age analysis controlling for never smoking status and other variables. Never smoking was not associated with COVID-19 status. The 50 to 69-year olds had a higher risk of testing positive for COVID-19. Besides that, males had a higher risk of testing positive for COVID-19, while patients with CHF history, ESRD, HIV positive and non-COVID-19 respiratory viral illness patients had a lower risk of testing positive for COVID-19. A complete analysis of COVID-19 and age is shown in Supplementary 1 tables S4 and S5.

**Table 1:**
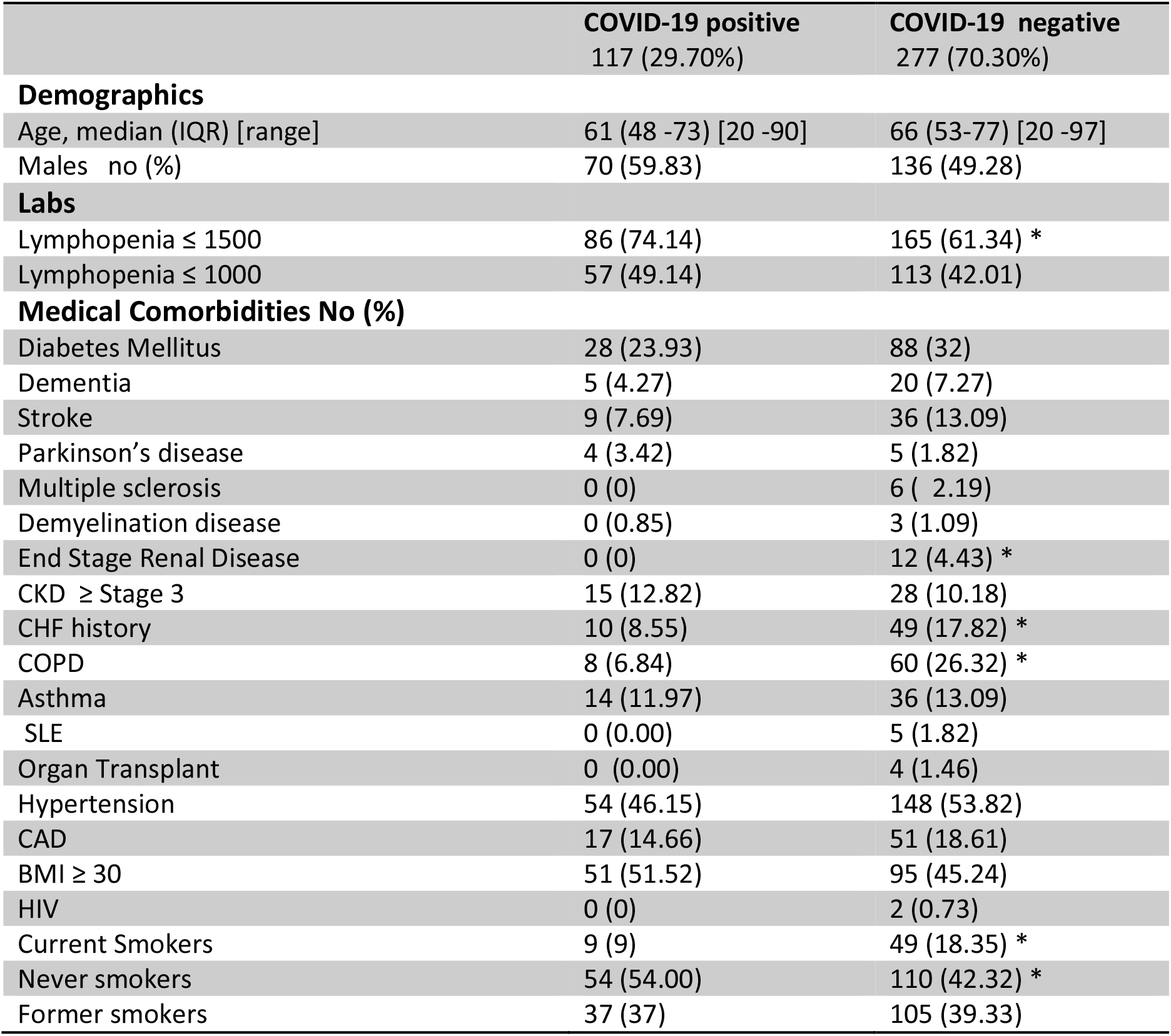
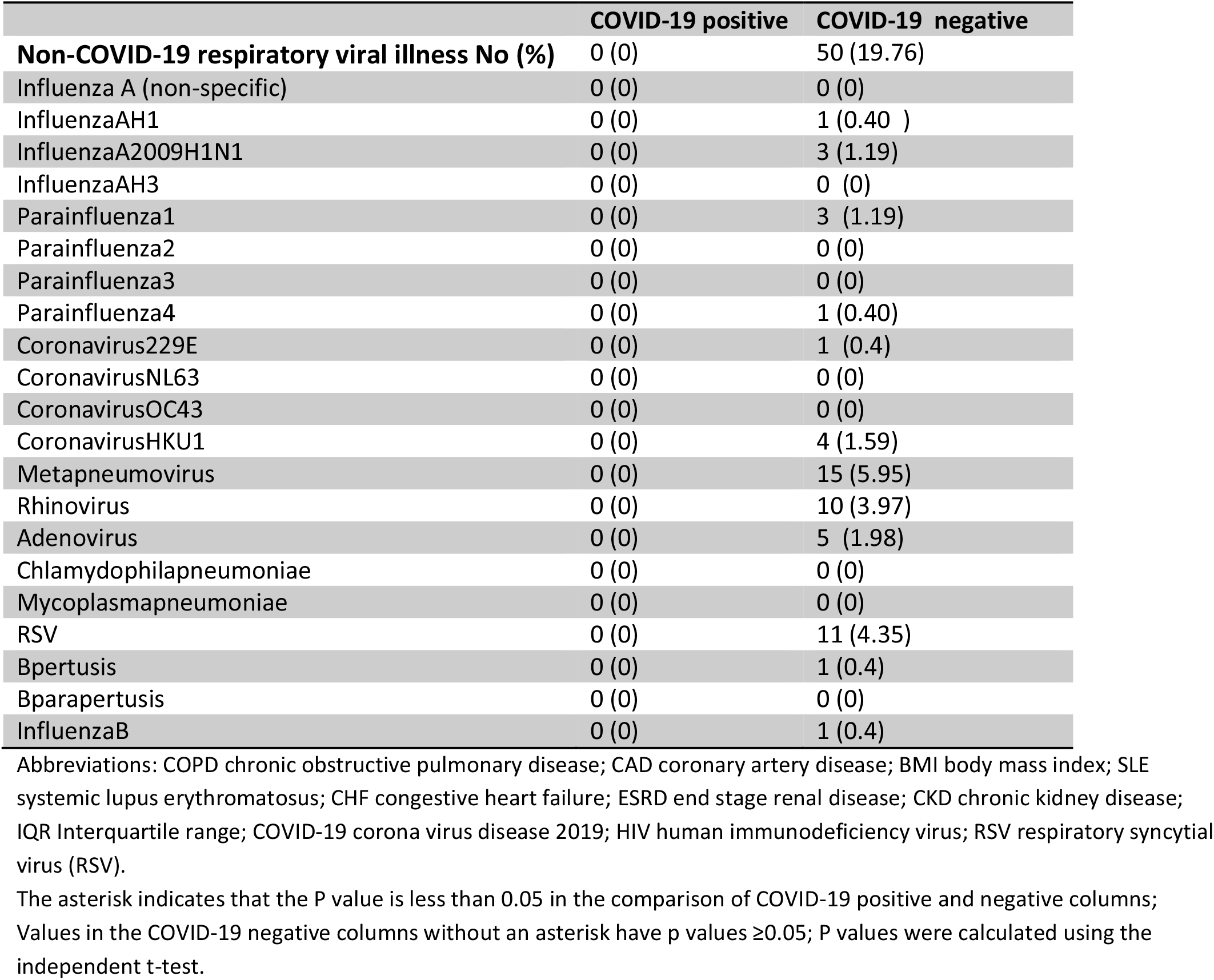
Baseline characteristics of study enrollees.

**Table 2:**
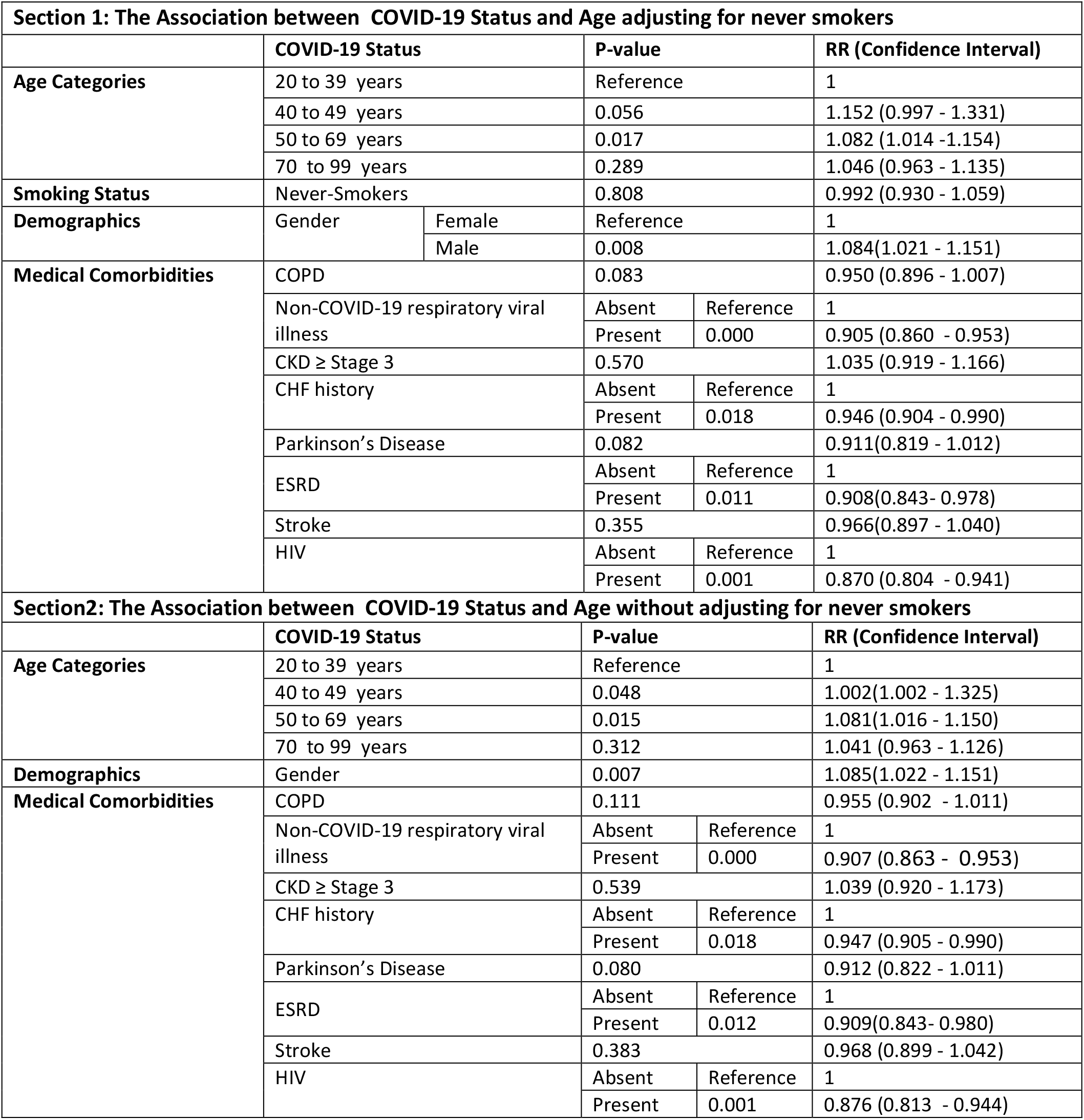
An analysis of COVID-19 status and age controlling for never smoking.

Section 2 of table 2 above shows partial results of the association between COVID-19 status and age without adjusting for never smoking. The 50 to 69-year olds and males were more likely to test positive for COVID-19, while CHF history, ESRD, HIV positive and non-COVID-19 respiratory viral illness patients were less likely to test positive for COVID-19.

### Is never smoking a confounder?

The analysis in tables 2 above shows that the association between COVID-19 status and age is not confounded by never smoking. Similarly, never smoking doesn’t confound the association between COVID-19 status and Gender, CHF, HIV or non-COVID-19 respiratory viral illness.

### Stratified analysis based on Never Smoking status

Section 1 of table 3 below shows partial results for the analysis of COVID-19 status and age among those with history of smoking (current and former smokers). The 40 to 49-year olds were more likely to test positive for COVID-19. While the HIV positive and non-COVID-19 respiratory viral illness patients were less likely to test positive for COVID-19. Please find the complete stratified analysis results of COVID-19 and age in Supplementary 1 tables S6 and S7.

**Table 3:**
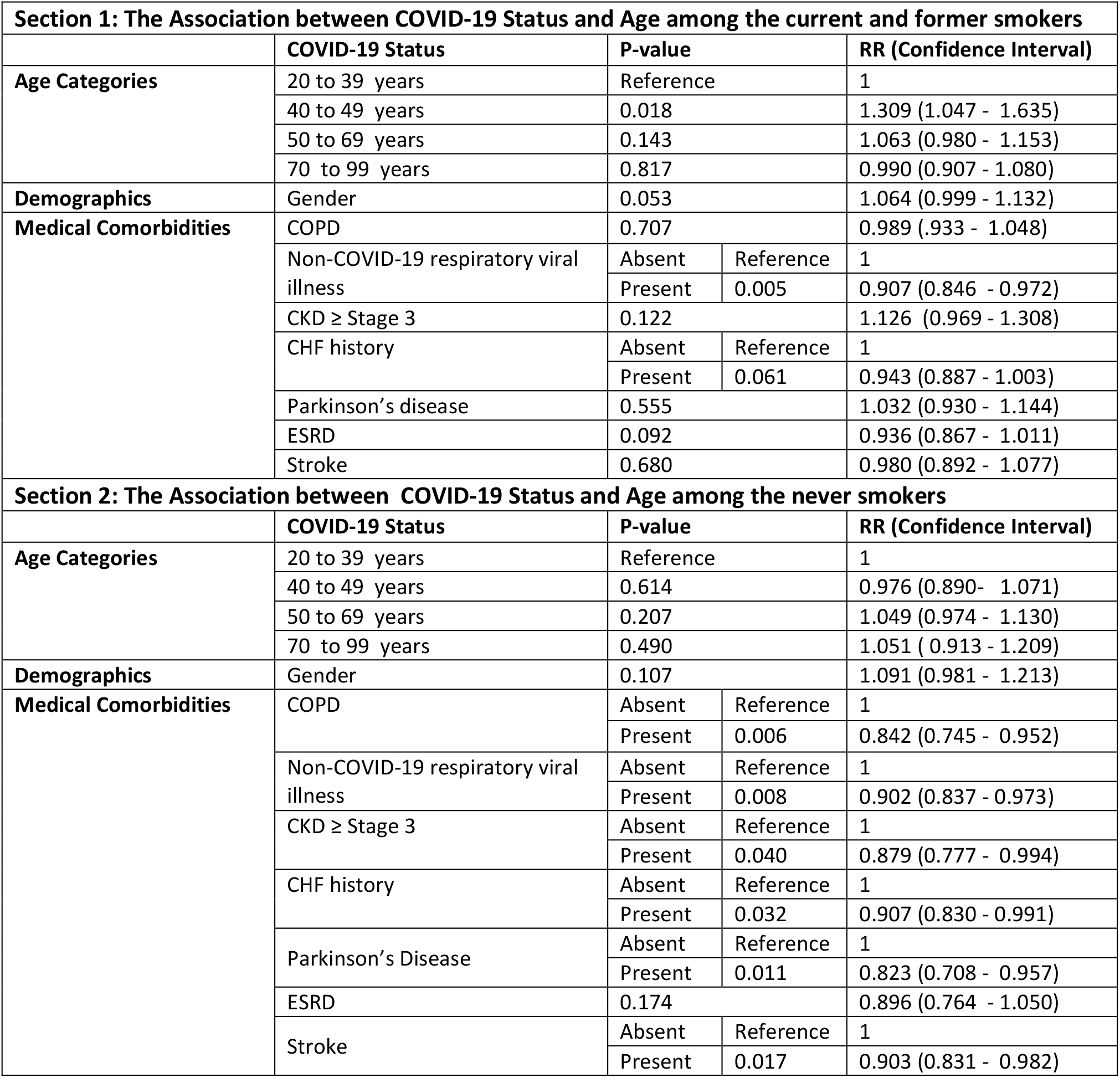
Stratified analysis based on Never Smoking status.

Section 2 of table 3 above shows partial results of COVID-19 status and age association among the never smokers. Age was not significantly associated with COVID-19 status, although 40 to 49-year old patients were less likely to test positive for COVID-19. Stroke, COPD, CHF history, CKD, Parkinson’s disease and non-COVID-19 respiratory viral illness patients were less likely to test positive for COVID-19.

The difference between the never smokers and those with history of smoking (current plus former smokers) among COPD, CHF, Parkinson disease and the 40-49-year-old patients was significantly different as shown by the homogeneity test in Supplementary 1 table S3.

The COVID-19 and age analysis among the never smoker and current smokers (excluding former smokers) shown in supplementary 2 tables, generated similar results to the analysis of never smokers versus those with history of smoking (current smokers +former smokers) presented above.

## Discussion

We found that 40-49-year-old current and former smokers were more likely to test positive for COVID-19. Although males and the 50-69-year-old patients were at higher risk of testing positive for COVID-19, smoking status had no influence on their COVID-19 positivity risk.

CHF, COPD and Parkinson disease never smokers were less likely to test positive for COVID-19. On the other hand, non-COVID-19 respiratory viral illness and ESRD disease patients had a lower risk of testing positive for COVID-19 regardless of their smoking status.

### What we know about COVID-19 and Smoking

To our knowledge, this is the first study to show that smoking increases the risk of testing positive for COVID-19, while never smoking reduces the risk of COVID-19 positivity.

The association between COVID-19 and smoking has attracted a lot interest,^8,9,10,11^ with some speculating that smoking reduces the risk of COVID-19 infection, given that published research has found a high prevalence of never smokers and a low prevalence of current smokers among COVID-19 positive patients. ^1,2^,^3^ However, these studies never compared COVID-19 positive and negative patients.

According to the WHO scientific brief on smoking and COVID-19 last updated on June 30^th^ 2020, “there are no peer-reviewed articles assessing the risk of COVID-19 among smokers”.^12^ There are 3 yet to be peer reviewed publications conducted in Israel, United Kingdom (UK) and France assessing smoking and COVID-19 status.^13,14, 15^

The Israel study found that never smokers had a higher risk of testing positive for COVID-19 compared to current and former smokers, while the UK study found no association between smoking and COVID-19 positivity.^13,14^ On the other hand, the France study found that COVID-19 positive patients on nicotine replacement therapy without smoking related diseases (all types of cancer, cardiovascular and respiratory diseases) were less likely to be hospitalized, whereas nicotine replacement therapy was not associated with the risk of hospitalization among COVID-19 positive patients with smoking related diseases.^15^

Sarah et al found the odds of current-smokers self-reporting to have been diagnosed with COVID-19 higher than never-smokers, in a UK online and telephone survey examining COVID-19 mental and social impact;^16^ it is unclear whether participants had a COVID-19 PCR test. ^16^

Despite the risk of testing positive for COVID-19 among smokers being unclear, current smokers and former smokers have a higher risk of morbidity, mortality and need for mechanical ventilation during hospitalization compared to COVID-19 never smokers.^2^ On the other hand, countries with the lowest prevalence of smoking have the highest COVID-19 mortality, while countries with the highest smoking prevalence have the lowest COVID-19 mortality according to Norden et al.^17^

### Strengths of this study

Our study compared patients who tested positive and negative for COVID-19 during hospitalization and stratified them based on their smoking status [never smokers versus history of smoking (current + former smokers)]. Moreover, the smoking status data was collected at the time of COVID-19 testing in our study.

Although the Israel study compared COVID-19 positive and negative patients, they didn’t stratify patients based on their smoking status.^13^ In addition, the smoking status in the Israel study was collected during prior medical visits before the COVID-19 pandemic.

Israel et al conducted a case controlled study using data of patients who sought (out-patient and in-patient) care from the Israel health care network.^13^ Each COVID-19 positive patient was matched to five COVID-19 negative patients of the same sex, age and ethnicity or religion.^13^ Most of the enrollees in the Israel study were not hospitalized, while most of our study enrollees were hospitalized. ^13^ Israel et al found that never-smokers were more likely to test positive for COVID-19 compared to current and former smokers, in contrast to our study findings.^13^

Cho et al^14^ in the UK study compared COVID-19 positive and negative patients enrolled in a large prospective study between 2006 and 2010, aimed at studying genetics, environment and lifestyle determinants in UK; ^14^ in contrast, our study examined hospitalized patients in USA. The smoking status data in the UK study was collected during enrollment between 2006 to 2010.^14^ Cho et al study stratified patients based on gender and excluded patients with respiratory disease; ^14^ while our study stratified patients based on their smoking status and included patients with respiratory disease. Consistent with the non-stratified results of our study, Cho et al found that current and past smoking was not associated with testing positive for COVID-19 compared to never smoking. ^14^

The France study compared people who were reimbursed and those not reimbursed for nicotine replacement therapy. Nicotine therapy reimbursed patients were assumed to have been current-smokers. However, nicotine replacement therapies are prescribed to patients who quit smoking to prevent nicotine craving and smoking relapse. ^15^ Therefore, patients who were not reimbursed for nicotine replacement therapy could have been current-smokers, never-smokers or former-smokers denied nicotine replacement reimbursement.

### Prevalence of smoking

The WHO brief reported that 1.4 – 18.5% of all hospitalized patients were current smokers.^12^ In consistent with the WHO brief, our study found that 15.76% of our study enrollees were current smokers.

The UK and Israel studies examined non-hospitalized (majority) and hospitalized enrollees. ^13,14^ The UK study (16.6%) had a higher prevalence of COVID-19 positive currents smokers compared to the Israel study (9.8%).^13,14^ Similarly the UK study (26.95%) had a higher prevalence of COVID-19 positive former smokers in contrast to the Israel (11.6%) study. ^13,14^ On the other hand, the Israel study (78.5%) had a higher prevalence of COVID-19 positive never smokers compared to the UK study (52.6%).^13, 14^

The prevalence of current smokers among COVID-19 positive patients was similar in the Cleveland study (8.6%) and our study (8.91%) during hospitalization.^3^ On the other hand, the mainland China and the New York, USA studies reported a 12.6% and 5.1% current smoking prevalence respectively among hospitalized COVID-19 positive patients.^1,2^

The mainland china (1.9%) study had the lowest prevalence of COVID-19 positive former smokers compared to the Cleveland clinic (36.4%), New York study (19.9%) and our study (37%).^2,3^,^4^The prevalence of COVID-19 positive never smokers was similar in the New York (84.4%) and the mainland China (85.4%) studies,^1,2^ while our study and the Cleveland Clinic USA study had a low and similar never smokers’ prevalence of 54% and 52 % respectively for COVID-19 positive patients^3^.

### Age and Smoking

Studies with a higher median and interquartile age [our study 61 years (48 -73 years) and the Cleveland clinic 64.37 years (54.83 – 76.58 years) study]^3^ had a lower prevalence of never smokers and a higher prevalence of former smokers, while studies with a lower median and interquartile age [the Mainland China 47 years (35 -58 years) and the Israel 39.54 years (27.38-58.26 years) studies] tended to have a higher never smokers’ prevalence and a low former smokers’ prevalence.^2,13^

Contrary to the aforementioned trend, the Richardson et al and Goyal et al New York studies with a median and interquartile age of [63 years (52-75 years)] and [62.2 years (48.6 -73.7 years)] respectively had a high never smokers’ prevalence of 75 and 84% respectively,^1,4^ while the Seattle study with a median age of 69 years also had a high never smokers’ prevalence of 74%.^6^ The UK study neither reported the median nor the interquartile age.^14^

Studies with a higher prevalence of never smokers had a low prevalence of former smokers and vice versa, since the current smoker’s prevalence (5.1% to 12.6 %) varied minimally irrespective of the median and the interquartile age of the COVID-19 positive enrollees. According to the CDC, the 25 to 44 year olds and 45 to 64-year olds have the highest current smoking prevalence of 16.5% and 16.3% respectively in the USA general population.^18^ While the current smoking prevalence among the 18 to 24 year olds and those ≥ 65-years old is 7.8% and 8.4% respectively.^18^

On the other hand, our study found that the 70-to 100-year-old patients had the lowest prevalence of current smokers (4.79%) while all the other age groups [(20-39-years), (40–49-years) and (50-69-years)] had a similar current smokers’ prevalence of approximately 23%.

Our study found that 40-49-year-old current and former smokers had a higher risk of testing positive for COVID-19 compared to the 20-to-39-year-old current and former smokers, despite both age groups having a similar current smokers’ prevalence. However, the 40-to-49-year-olds had a lower former smokers’ prevalence; perhaps the lower former-smokers’ prevalence increased the 40-49-year olds risk of COVID-19 positivity. On the other hand, the 40-49-year-old never smokers had a lower COVID-19 positivity risk, though not significant statistically. Moreover, the 50-69-year-old patients were more likely to test positive for COVID-19 regardless of their smoking status.

### Medical comorbidities

The Israel et al study found that obese patients were more likely to test positive for COVID-19 [1.172(1.084-1.267)] while patients with hypertension [0.834(0.738-0.941)], asthma [0.766(0.654-0.896)] and malignancy [0.788(0.677-0.917)] were less likely to test positive for COVID-19.^13^ However, the Israel study used BMI calculated from weight and height collected prior to the COVID-19 pandemic.^13^

The UK study found that > 80 years old men [1.92(1.08-3.41)], men with a BMI of 27-<30 [1.64 (1.12-2.39)] and men with a BMI of ≥30, [1.60(1.10-2.32)] were more likely to test positive for COVID-19.^14^ However, the BMI in the UK study was calculated from weight and height collected during enrollment between 2006 -2010.^14^ In contrast, our study found that BMI and the 70 to 100-year olds were not associated with testing positive for COVID-19. Moreover, the UK study didn’t control for COPD because they excluded patients with respiratory disease.^14^

Our study found that CHF history, COPD and Parkinson disease never smokers had a lower risk of testing positive for COVID-19, while ESRD and non-COVID-19 respiratory viral illness patients had a lower risk of testing positive for COVID-19.

### Pathogenesis of COVID-19 among smokers

It has been hypothesized that the COVID-19 virus attaches to the angiotensin converting enzyme 2 (ACE-2) receptors using the spike (S) glycoprotein during pulmonary epithelial cell entry.^19^ Current smokers, former smokers and COPD patients are more likely to have an increased ACE-2 expression in their small airways^20,21,22^

Our study found that, although 40-49-year-old former and current smokers were at higher risk of testing positive for COVID-19 than the 20-39-year olds, the prevalence of current smokers was not significantly different between the 40 to 49-year olds and the 20 to 39-year olds. On the other hand, COPD never smokers were less likely to test positive for COVID-19, while COPD current and former smokers were neither at a higher risk nor at lower risk of testing positive for COVID-19.

Our study suggests that having an increased expression of ACE-2 receptors alone doesn’t increase the risk of testing positive for COVID-19, given that that current smokers, former smokers and COPD patients have an increased ACE-2 expression.^20^

### Limitations

Limitations of our study included missing data and not all patients had laboratory tests done. Even though we didn’t have data on how long the former smokers had quit smoking, the analysis of never smokers versus current smokers (excluding former smokers) shown in supplementary S2 yielded similar results.

## Conclusion

Based on our study findings, smoking increases the risk of testing positive for COVID-19 among the 40 to 49-year-old current and former smokers, while never smoking reduces the risk of testing positive for COVID-19 among the CHF, COPD and Parkinson disease patients.

Regardless of their smoking status, males and the 50-69-year-olds are more likely to test positive for COVID-19, while ESRD and non-COVID-19 respiratory viral illness patients are less likely to test positive for COVID-19.

Our study emphasizes that smoking doesn’t reduce the risk of testing positive for COVID-19, while never smoking doesn’t increase the risk of COVID-19 positivity, despite COVID-19 positive patients having a low current-smokers’ prevalence and a high never-smokers’ prevalence.

## Supporting information

Supplementary 1 will be used for the link on the preprint site

Supplementary 2 will be used for the link to the file on the preprint site

## Data Availability

all the data is available in the manuscript and in the supplementary files

## Acknowledgements

We want to thank the following groups for their support: The COVID-19 incident command Center PeaceHealth Sacred Heart Medical Center at Riverbend; The PeaceHealth Tableau Developers; the PeaceHealth Sacred Heart Medical Center at Riverbend and The Hospitalists group Sacred Heart Medical Center at Riverbend. We also thank Amy Ballard and Antony Wala.

## Authors Contributions

Samson Barasa: concept, design, data acquisition, analysis and interpretation, Preparation of manuscript

Josephine Kiage-Mokaya: data acquisition and preparation of manuscript Katya Cruz-Madrid: Preparation of manuscript

Michael Friedlander: data acquisition and Preparation of manuscript

## SUPPORTING INFORMATION

Additional supporting information is in Supplementary 1 and 2.

**Supplementary 1:** The analysis of COVID-19 and Age among the never-smokers versus current-smokers plus former-smokers)

Table S1: Baseline characteristics of never smokers

Table S2: Baseline characteristics of those with history of smoking (current and former smokers)

Table S3: The homogeneity test

Table S4: The Association between COVID-19 Status and Age adjusting for never smokers

Table S5: The Association between COVID-19 Status and Age with adjusting for never smokers

Table S6: The Association between COVID-19 Status and Age among current and former smokers

Table S7: The Association between COVID-19 Status and Age among never smokers **Abbreviations:**

**Supplementary 2:** The analysis of COVID-19 and Age among the never-smokers versus current-smokers (excluding the former-smokers)

Table T1: The Association between COVID-19 Status and Age adjusting for smoking status

Table T2: The Association between COVID-19 Status and Age without adjusting for Smoking status

Table T3: The Association between COVID-19 Status and Age among the never smokers

Table T4: The Association between COVID-19 Status and Age among the current smokers

Table T5: The homogeneity test **Abbreviations:**

## References

1. Richardson S, Hirsch JS, Narasimhan M, et al. Presenting Characteristics, Comorbidities, and Outcomes Among 5700 Patients Hospitalized With COVID-19 in the New York City Area. JAMA : the journal of the American Medical Association. 2020.

2. Ding Q, Lu P, Fan Y, et al. The clinical characteristics of pneumonia patients co-infected with 2019 novel coronavirus and influenza virus in Wuhan, China. Journal of medical virology. 2020.

3. Jehi L, Ji X, Milinovich A, et al. Development and validation of a model for individualized prediction of hospitalization risk in 4,536 patients with COVID-19. PloS one. 2020;15(8):e0237419.

4. Goyal P, Choi JJ, Pinheiro LC, et al. Clinical Characteristics of Covid-19 in New York City. The New England journal of medicine. 2020;382(24):2372–2374.

5. Jehi L, Ji X, Milinovich A, et al. Individualizing Risk Prediction for Positive Coronavirus Disease 2019 Testing: Results from 11,672 Patients. Chest. 2020.

6. Buckner FS, McCulloch DJ, Atluri V, et al. Clinical Features and Outcomes of 105 Hospitalized patients with COVID-19 in Seattle, Washington. Clinical infectious diseases : an official publication of the Infectious Diseases Society of America. 2020.

7. Barasa S, Kiage-Mokaya J, Luna G, et al. The major predictors of testing positive for COVID-19 among symptomatic hospitalized patients. medRxiv preprint. 2020.

8. Crist C. Smokers Hospitalized Less Often for COVID-19 Available: www.webmd.com/lung/news/20200430/smokers-hospitalized-less-often-for-covid-19.. Accessed 20 Nov. 2020, WebMD.

9. Robertson S. An inverse relationship between smoking and COVID-19 Available: https://www.news-medical.net/news/20200615/An-inverse-relationship-between-smoking-and-COVID-19.aspx. Accessed 20 Nov. 2020, 2020.

10. Robertson S. Exploring the effects of smoking tobacco on COVID-19 risk Available: https://www.news-medical.net/news/20201014/Exploring-the-effects-of-smoking-tobacco-on-COVID-19-risk.aspx. Accessed 20 Nov. 2020, 2020.

11. Boyd C. What do we actually know about smoking and COVID-19? Available: https://www.rstreet.org/2020/08/04/what-do-we-actually-know-about-smoking-and-covid-19/. Accessed 11/20/2020, 2020.

12. WHO. Smoking and COVID-19 Available: https://www.who.int/news-room/commentaries/detail/smoking-and-covid-19. Accessed 9/27/20, 2020.

13. Israel A, Felhamer E, Lahad A, et al. Smoking and the risk of COVID-19 in a large observational population study. medRxiv preprint. 2020.

14. Cho ER, Jha P, Slusky AS. Smoking and the risk of COVID-19 infection in the UK Biobank Prospective medRxiv preprint. 2020.

15. Mahmoud Zureik MD, Ph.D.,, Bérangère Baricault, Clémentine Vabre PD, et al. Nicotine-replacement therapy, as a surrogate of smoking, and the risk of hospitalization with Covid-19 and all-cause mortality: a nationwide, observational cohort study in France. medRxiv preprint. 2020.

16. Jackson SE, Brown J, Shahab L, et al. COVID-19, smoking and inequalities: a study of 53 002 adults in the UK. Tobacco control. 2020.

17. Norden MJ, Avery DH, Norden JG, et al. National Smoking Rates Correlate Inversely with COVID-19 Mortality. medRxiv preprint. 2020.

18. CDC. Current Cigarette Smoking Among Adults in the United States Available: https://www.cdc.gov/tobacco/data_statistics/fact_sheets/adult_data/cig_smoking/index.htm. Accessed 9/26/2020, 2020.

19. Perrotta F, Matera MG, Cazzola M, et al. Severe respiratory SARS-CoV2 infection: Does ACE2 receptor matter? Respiratory medicine. 2020;168:105996.

20. Leung JM, Yang CX, Tam A, et al. ACE-2 expression in the small airway epithelia of smokers and COPD patients: implications for COVID-19. The European respiratory journal. 2020;55(5).

21. Cai G, Bosse Y, Xiao F, et al. Tobacco Smoking Increases the Lung Gene Expression of ACE2, the Receptor of SARS-CoV-2. American journal of respiratory and critical care medicine. 2020;201(12):1557–1559.

22. Polverino F. Cigarette Smoking and COVID-19: A Complex Interaction. American journal of respiratory and critical care medicine. 2020;202(3):471–472.

