## Supplementary 1 will be used for the link on the preprint site for "Smoking increases the risk of COVID-19 positivity, while Never-smoking reduces the risk"

**The analysis of COVID-19 and Age among the never smokers and those with history of smoking (current smokers and former smokers)**

### Supplementary 1: Table S1

**Table S1: Baseline characteristics of never smokers**

| | **COVID-19 positive** 71 (36.60%) | **COVID-19 Negative** 123 (63.4 %) |
|---|---|---|
| **Demographics** | | |
| Age, median (IQR) [range] | 60 (47 -70) [20 - 90] | 65 (48-78) [24-97] |
| Males no (%) | 37 (52.11) | 58 (47.54) |
| **Labs** | | |
| Lymphopenia ≤ 1500 | 52 (74.29) | 71 (60.17) |
| Lymphopenia ≤ 1000 | 38 (54.29) | 43 (36.44) * |
| **Medical Comorbidities No (%)** | | |
| Diabetes Mellitus | 14 (19.72) | 38 (31.15) |
| Dementia | 1 (1.41) | 7 (5.74) |
| Stroke | 5 (7.04) | 12 (9.84) |
| Parkinson's | 1 (1.41) | 3 (2.46) |
| Multiple sclerosis | 0 (0) | 1 (0.82) |
| Demyelinating Disease | 1 (1.41) | 0 (0) |
| End Stage Renal Disease | 0 (0) | 5 (4.13) |
| CKD ≥ Stage 3 | 3 (4.23) | 9 (7.38) |
| CHF history | 3 (4.23) | 17 (13.93) * |
| COPD | 1 (1.41) | 7 (5.74) |
| Asthma | 7 (9.86) | 17 (13.93) |
| SLE | 0 (0) | 2 (1.65) |
| Organ Transplant | 0 (0) | 3 (2.46) |
| Hypertension | 27 (38.03) | 58 (47.54) |
| CAD | 6 (8.57) | 17 (14.05) |
| BMI ≥ 30 | 28 (48.28) | 45 (45.45) |
| HIV | 0 (0.00) | 0 (0.00) |
| Non-COVID-19 respiratory viral illness | 0 (0) | 17 (15.6) |

Abbreviations: COPD chronic obstructive pulmonary disease; CAD coronary artery disease; BMI body mass index; SLE systemic lupus erythromatosus; CHF congestive heart failure; CKD chronic kidney disease; IQR Interquartile range; COVID-19 corona virus disease 2019 ; HIV human immunodeficiency virus.

The asterisk indicates that the P value is less than 0.05 in the comparison of COVID-19 positive and negative column; Values in the COVID-19 negative columns without an asterisk have p values ≥0.05; P values were calculated using the independent t-test.

### Supplementary 1: Table S2

**Table S2: Baseline characteristics of those with history of smoking (current and former smokers)**

| | **COVID-19 positive** 63 (27.75%) | **COVID-19 Negative** 164 (72.25 %) |
|---|---|---|
| **Demographics** | | |
| Age, median (IQR) [range] | 61 (44 -76) [20 - 88] | 66 (55-76) [20-97] |
| Males no (%) | 43 (68.25) | 83 (50.61) * |
| **Labs** | | |
| Lymphopenia ≤ 1500 | 46 (73.02) | 101 (62.73) |
| Lymphopenia ≤ 1000 | 28 (44.44) | 77 (47.83) |
| **Medical Comorbidities No (%)** | | |
| Diabetes Mellitus | 16 (25.40) | 51 (31.29) |
| Dementia | 4 (6.35) | 13 ( 7.98) |
| Stroke | 5 (7.94) | 24 (14.72) |
| Parkinson's | 3 (4.76) | 2 (1.23) |
| Multiple sclerosis | 0 (0) | 6 (3.70) |
| Demyelination Disease | 1 (1.59) | 3 (1.84) |
| End Stage Renal Disease | 0 (0) | 7 ( 4.38) |
| CKD ≥ Stage 3 | 12 (19.05) | 19 (11.66) |
| CHF history | 7 (11.11) | 32 (19.63) |
| COPD | 8 (12.70) | 61 (37.42) * |
| Asthma | 8 (12.70) | 19 (11.66) |
| SLE | 0 (0) | 4 (2.45) |
| Organ Transplant | 0 (0) | 1 (0.62) |
| Hypertension | 29 (46.03) | 93 (57.06) |
| CAD | 11 (17.46) | 35 (21.47) |
| BMI ≥ 30 | 27 (51.92) | 66 (46.48) |
| HIV | 0 (0) | 2 (1.23) |
| Non-COVID-19 respiratory viral Illness | 0 (0) | 32 (21.33) |

Abbreviations: COPD chronic obstructive pulmonary disease; CAD coronary artery disease; BMI body mass index; SLE systemic lupus erythromatosus; CHF congestive heart failure; CKD chronic kidney disease; IQR Interquartile range; COVID-19 corona virus disease 2019 ; HIV human immunodeficiency virus.

The asterisk indicates that the P value is less than 0.05 in the comparison of COVID-19 positive and negative column; Values in the COVID-19 negative columns without an asterisk have p values ≥0.05; P values were calculated using the independent t-test.

### Supplementary 1: Table S3

Table S3 below shows homogeneity test. The difference between never smokers and those with history of smoking (current plus former smokers) among the COPD, CHF, Parkinson disease and the 40 to 49-year olds is significantly different.

| **Table S3: The homogeneity test** | | | | |
|---|---|---|---|---|
| | **COVID-19 Status** | **P-Value** | | **RR (Confidence Interval)** |
| **Smoking Status** | Never Smokers | 0.524 | | 1.023 (0.954-1.097) |
| **Demographics** | **Age Categories** | 20 | Reference | 1 |
| | | 40 | 0.027 | 1.292 (1.030-1.620) |
| | | 50 | 0.048 | 1.089 (1.001-1.184) |
| | | 70 | 0.343 | 1.040 (0.959-1.128) |
| | AgeCat6#NeverSmokers | 20 2 | Reference | 1 |
| | | 40 2 | 0.034 | 0.779 (0.618-0.982) |
| | | 50 2 | 0.740 | 0.980 (0.872-1.102) |
| | | 70 2 | 0.734 | 1.021 (0.904-1.154) |
| | Gender | 0.018 | | 1.074 (1.012-1.140) |
| **Medical comorbidities** | COPD#c.NeverSmokers | No | Reference | 1 |
| | | Yes | 0.018 | 0.950 (0.910-0.991) |
| | CHF#c.NeverSmokers | No | Reference | 1 |
| | | Yes | 0.009 | 0.958 (0.928-0.989) |
| | Diabetes | 0.712 | | 0.987 (0.921-1.058) |
| | Hypertension | 0.068 | | 1.067 (0.995-1.143) |
| | Non-COVID-19 respiratory viral illness | 0.000 | | 0.910 (0.866 - 0.955) |
| | BMI4 | 0.136 | | 1.004 (0.999-1.008) |
| | Lymphocyt1e | 0.525 | | 1.020 (0.961-1.082) |
| | Asthma | 0.354 | | 1.054 (0.943-1.179) |
| | CKD#c.NeverSmokers | No | Reference | 1 |
| | | Yes | 0.902 | 0.995 (0.923-1.073) |
| | SLE | 0.119 | | 0.891 (0.771-1.030) |
| | Organ Transplant | 0.107 | | 0.898 (0.789-1.023) |
| | CAD | 0.690 | | 1.017 (0.936-1.105) |
| | HIV | 0.001 | | 0.860 (0.785-0.943) |
| | Parkinsons#c.NeverSmoker | No | Reference | 1 |
| | | Yes | 0.023 | 0.935 (0.883-0.991) |
| | Multiple sclerosis | 0.078 | | 0.912 (0.823-1.011) |
| | ESRD | 0.024 | | 0.916 (0.849-0.989) |
| | Stroke#c.NeverSmoker | No | Reference | 1 |
| | | Yes | 0.229 | 0.972 (0.927-1.018) |
| | Demyelinatingdisease | 0.291 | | 0.929 (0.809-1.066) |

### Supplementary 1: Table S4

Table S4 below shows the results of the association between COVID-19 status and age without adjusting for never smoking. The 50 to 69-year olds had a higher risk of testing positive for COVID-19. Besides that, males had a higher risk of testing positive for COVID-19, while patients with CHF history, ESRD, HIV positive and non-COVID-19 respiratory viral illness patients had a lower risk of testing positive for COVID-19.

**Table S4: The Association between COVID-19 Status and Age adjusting for never smokers**

| | **COVID-19 Status** | | **P-value** | | **RR (Confidence Interval)** |
|---|---|---|---|---|---|
| **Age Categories** | 20 to 39 years | | Reference | | 1 |
| | 40 to 49 years | | 0.056 | | 1.152 (0.997 - 1.331) |
| | 50 to 69 years | | 0.017 | | 1.082 (1.014 -1.154) |
| | 70 to 99 years | | 0.289 | | 1.046 (0.963 - 1.135) |
| **Smoking Status** | Never Smokers | | 0.808 | | 0.992 (0.930 - 1.059) |
| **Demographics** | Gender | Female | Reference | | 1 |
| | | Male | 0.008 | | 1.084(1.021 - 1.151) |
| **Labs** | Lymphopenia (≤1500) | | 0.757 | | 1.009 (.951 - .0715) |
| **Medical Comorbidities** | COPD | | 0.083 | | 0.950 (0.896 - 1.007) |
| | Non-COVID-19 respiratory viral illness | | Absent | Reference | 1 |
| | | | Present | 0.000 | 0.905 (0.860 - 0.953) |
| | Hypertension | | 0.109 | | 1.061 (0.987 - 1.141) |
| | Diabetes | | 0.573 | | 0.980 (0.915 - 1.051) |
| | Asthma | | 0.422 | | 1.047 (0.936 - 1.171) |
| | SLE | | 0.301 | | 0.945 (0.848 - 1.052) |
| | Organ Transplant | | 0.090 | | 0.898 (0.792 - 1.017) |
| | CKD ≥ Stage 3 | | 0.570 | | 1.035 (0.919 - 1.166) |
| | CHF history | | Absent | Reference | 1 |
| | | | Present | 0.018 | 0.946 (0.904 - 0.990) |
| | CAD | | 0.726 | | 1.015(0.935-1.101) |
| | Parkinson’s Disease | | 0.082 | | 0.911(0.819 - 1.012) |
| | Multiple sclerosis | | 0.183 | | 0.946(0.872- 1.027) |
| | ESRD | | Absent | Reference | 1 |
| | | | Present | 0.011 | 0.908(0.843- 0.978) |
| | Stroke | | 0.355 | | 0.966(0.897 - 1.040) |
| | Demyelinating disease | | 0.296 | | 0.930(0.811 - 1.066) |
| | HIV | | Absent | Reference | 1 |
| | | | Present | 0.001 | 0.870 (0.804 - 0.941) |
| | BMI ≥ 30 | | 0.193 | | 1.003 (0.998 - 1.008) |

### Supplementary 1: Table S5

Table S5 below shows the results of the association between COVID-19 status and age without adjusting for never smoking. The 50 to 69-year olds and males were more likely to test positive for COVID-19, while CHF history, ESRD, HIV positive and patients with non-COVID-19 respiratory viral illness were less likely to test positive for COVID-19.

| **Table S5: The Association between COVID-19 Status and Age with adjusting for never smokers** | | | | |
|---|---|---|---|---|
| | **COVID-19 Status** | **P-value** | | **RR (Confidence Interval)** |
| **Age Categories** | 20 to 39 years | Reference | | 1 |
| | 40 to 49 years | 0.048 | | 1.002(1.002 - 1.325) |
| | 50 to 69 years | 0.015 | | 1.081(1.016 - 1.150) |
| | 70 to 99 years | 0.312 | | 1.041 (0.963 - 1.126) |
| **Demographics** | Gender | 0.007 | | 1.085(1.022 - 1.151) |
| **Labs** | Lymphopenia (≤1500) | 0.771 | | 1.009 (0.951 - 0.070) |
| **Medical Comorbidities** | COPD | 0.111 | | 0.955 (0.902 - 1.011) |
| | Non-COVID-19 respiratory viral illness | Absent | Reference | 1 |
| | | Present | 0.000 | 0.907 (0.863 - 0.953) |
| | Hypertension | 0.095 | | 1.062 (0.989 - 1.140) |
| | Diabetes | 0.568 | | 0.980 (0.916 - 1.049) |
| | Asthma | 0.411 | | 1.048 (0.937 - 1.172) |
| | SLE | 0.167 | | 0.945 (0.873 - 1.024) |
| | Organ Transplant | 0.063 | | 0.894 (0.793 - 1.006) |
| | CKD ≥ Stage 3 | 0.539 | | 1.039 (0.920 - 1.173) |
| | CHF history | Absent | Reference | 1 |
| | | Present | 0.018 | 0.947 (0.905 - 0.990) |
| | CAD | 0.711 | | 1.015(0.937-1.008) |
| | Parkinson’s Disease | 0.080 | | 0.912 (0.822 - 1.011) |
| | Multiple sclerosis | 0.148 | | 0.949 (0.884- 1.019) |
| | ESRD | Absent | Reference | 1 |
| | | Present | 0.012 | 0.909(0.843- 0.980) |
| | Stroke | 0.383 | | 0.968 (0.899 - 1.042) |
| | Demyelinating disease | 0.297 | | 0.932(0.817 - 1.064) |
| | HIV | Absent | Reference | 1 |
| | | Present | 0.001 | 0.876 (0.813 - 0.944) |
| | BMI ≥ 30 | 0.195 | | 1.003 (0.998 - 1.008) |

### Supplementary 1: Table S6

Table S6 below shows the analysis results the COVID-19 status and age among those with history of smoking (current and former smokers). The 40 to 49-year olds were more likely to test positive for COVID-19. While the HIV positive and non-COVID-19 respiratory viral illness patients were less likely to test positive for COVID-19.

**Table S6: The Association between COVID-19 Status and Age among current and former smokers**

| | **COVID-19 Status** | **P-value** | | **RR (Confidence Interval)** |
|---|---|---|---|---|
| **Age Categories** | 20 to 39 years | Reference | | 1 |
| | 40 to 49 years | 0.018 | | 1.309 (1.047 - 1.635) |
| | 50 to 69 years | 0.143 | | 1.063 (0.980 - 1.153) |
| | 70 to 99 years | 0.817 | | 0.990 (0.907 - 1.080) |
| **Demographics** | Gender | 0.053 | | 1.064 (0.999 - 1.132) |
| **Labs** | Lymphopenia (≤1500) | 0.270 | | 1.035 (0.974 - 1.100) |
| **Medical Comorbidities** | COPD | 0.707 | | .989 (.933 - 1.048) |
| | Non-COVID-19 respiratory viral illness | Absent | Reference | 1 |
| | | Present | 0.005 | 0.907 (0.846 - 0.972) |
| | Hypertension | 0.104 | | 1.080 (0.984 - 1.186) |
| | Diabetes | 0.150 | | 0.944 (0.874 - 1.021) |
| | Asthma | 0.758 | | 1.021 (.894 - 1.167) |
| | SLE | 0.188 | | 0.918 (0.808 - 1.043) |
| | Organ Transplant | Omitted | | 1 |
| | CKD ≥ Stage 3 | 0.122 | | 1.126 (0.969 - 1.308) |
| | CHF history | Absent | Reference | 1 |
| | | Present | 0.061 | 0.943 (0.887 - 1.003) |
| | CAD | 0.592 | | 1.023 (0.941 -1.113) |
| | Parkinson's disease | 0.555 | | 1.032 (0.930 - 1.144) |
| | Multiple sclerosis | 0.098 | | 0.898 (0.791 - 1.020) |
| | ESRD | 0.092 | | 0.936 (0.867 - 1.011) |
| | Stroke | 0.680 | | 0.980 (0.892 - 1.077) |
| | Demyelinating disease | 0.147 | | 0.929 (0.841 - 1.026) |
| | HIV | Absent | Reference | 1 |
| | | Present | 0.006 | 0.863 (0.775 - 0.959) |
| | BMI ≥ 30 | 0.359 | | 1.003 (.997 - 1.008) |

### Supplementary 1: Table S7

Table S7 below shows the results of the association between COVID-19 and age among the never smokers. Age was not associated with testing positive for COVID-19. Stroke, COPD, CHF history, CKD, Parkinson's disease and non-COVID-19 respiratory viral illness patients were less likely to test positive for COVID-19.

**Table S7: The Association between COVID-19 Status and Age among never smokers**

| | **COVID-19 Status** | **P-value** | | **RR (Confidence Interval)** |
|---|---|---|---|---|
| **Age Categories** | 20 to 39 years | Reference | | 1 |
| | 40 to 49 years | 0.614 | | 0.976 (0.890- 1.071) |
| | 50 to 69 years | 0.207 | | 1.049 (0.974 - 1.130) |
| | 70 to 99 years | 0.490 | | 1.051 ( 0.913 - 1.209) |
| **Demographics** | Gender | 0.107 | | 1.091 (0.981 - 1.213) |
| **Lab** | Lymphopenia (≤1500) | 0.795 | | 1.015 (0.906 - 1.137) |
| **Medical Comorbidities** | COPD | Absent | Reference | 1 |
| | | Present | 0.006 | 0.842 (0.745 - 0.952) |
| | Non-COVID-19 respiratory viral illness | Absent | Reference | 1 |
| | | Present | 0.008 | 0.902 (0.837 - 0.973) |
| | Hypertension | 0.212 | | 1.063 (0.966 - 1.171) |
| | Diabetes | 0.453 | | 1.040 (0.939 - 1.151) |
| | Asthma | 0.370 | | 1.087 (0.905 - 1.306) |
| | SLE | 0.252 | | 0.914 (0.785 - 1.065) |
| | Organ Transplant | 0.219 | | 0.906 (0.773 - 1.061) |
| | CKD ≥ Stage 3 | Absent | Reference | 1 |
| | | Present | 0.040 | 0.879 (0.777 - 0.994) |
| | CHF history | Absent | Reference | 1 |
| | | Present | 0.032 | 0.907 (0.830 - 0.991) |
| | CAD | 0.921 | | 1.007 (0.875 - 1.159) |
| | Parkinson's Disease | Absent | Reference | 1 |
| | | Present | 0.011 | 0.823 (0.708 - 0.957) |
| | Multiple sclerosis | | Omitted | 1 |
| | ESRD | 0.174 | | 0.896 (0.764 - 1.050) |
| | Stroke | Absent | Reference | 1 |
| | | Present | 0.017 | 0.903 (0.831 - 0.982) |
| | Demyelinating disease | Omitted | | 1 |
| | HIV | Omitted | | 1 |
| | BMI ≥ 30 | 0.541 | | 1.003 (0.995 - 1.010) |

### Abbreviations

ACE-2 angiotensin converting enzyme 2
CAD coronary artery disease
CDC center for disease control
COPD chronic obstructive pulmonary disease
BMI body mass index
SLE systemic lupus erythromatosus
CHF congestive heart failure
ESRD end stage renal disease
CKD chronic kidney disease
COVID-19 corona virus disease 2019
HIV human immunodeficiency virus
Interquartile range IQR
Polymerase chain reaction PCR
RSV respiratory syncytial virus
USA united states of America
UK United Kingdom
