## Supplementary 2 will be used for the link to the file on the preprint site for "Smoking increases the risk of COVID-19 positivity, while Never-smoking reduces the risk"

**The analysis of COVID-19 and Age among the never smokers and current smokers (excluding the former smokers)**

Table T1 below shows the association between COVID-19 and age adjusting for smoking status. The 50 to 69 year old patients were more likely to test positive for COVID-19, while COPD, Parkinson's disease and demyelinating disease patients were less likely to test positive for COVID-19.

| **Table T1: The Association between COVID-19 Status and Age adjusting for smoking status** | | | | |
|---|---|---|---|---|
| | COVID-19 | | P-Value | **RR (Confidence Interval)** |
| **Smoking status** | Never-Current Smokers | | 0.485 | 0.973 (0.901-1.051) |
| **Demographics** | Age Categories | 20-39 years | Reference | 1 |
| | | 40-49 years | 0.289 | 1.090 (0.930-1.278) |
| | | 50-69 years | 0.019 | 1.084 (1.013-1.159) |
| | | 70-100 years | 0.076 | 1.130 (0.987-1.293) |
| | Gender | | 0.060 | 1.081 (0.997-1.173) |
| **Medical comorbidities** | COPD | Absent | Reference | 1 |
| | | Present | 0.010 | 0.874 (0.789-0.968) |
| | CHF | | 0.096 | 0.943 (0.879-1.011) |
| | Diabetes | | 0.829 | 1.010 (0.925-1.102) |
| | Hypertension | | 0.396 | 1.037 (0.954-1.128) |
| | Non-COVID-19 respiratory viral illness | Absent | Reference | 1 |
| | | Present | 0.005 | 0.904 (0.842- 0.971) |
| | BMI4 | | 0.090 | 1.006 (0.999-1.014) |
| | Lymphocyte 1500 | | 0.874 | 0.994 (0.918-1.075) |
| | Asthma | | 0.565 | 1.035 (0.920-1.164) |
| | CKD | | 0.072 | 0.915 (0.831-1.008) |
| | SLE | | 0.759 | 0.968 (0.789-1.189) |
| | Organ Transplant | | 0.393 | 0.946 (0.834-1.074) |
| | CAD | | 0.501 | 0.967 (0.878-1.065) |
| | HIV | | | 1 |
| | Parkinson's disease | Absent | Reference | 1 |
| | | Present | 0.008 | 0.816 (0.702-0.905) |
| | Multiple sclerosis | | 0.812 | 0.983 (0.853-1.133) |
| | ESRD | | 0.212 | 0.938 (0.849-1.037) |
| | Stroke | | 0.211 | 0.956 (0.891-1.026) |
| | Demyelinating disease | Absent | Reference | 1 |
| | | Present | 0.036 | 0.860 (0.747-0.990) |
| | _cons | | 0.026 | 1.636 (1.06-2.523) |

Table T2 below shows the association between COVID-19 and age without adjusting for smoking status. The males and 50 to 69 year old patients were more likely to test positive for COVID-19, while CHF, HIV positive, ESRD and demyelinating disease patients were less likely to test positive for COVID-19.

| **Table T2: The Association between COVID-19 Status and Age without adjusting for Smoking status** | | | | |
|---|---|---|---|---|
| | COVID-19 | | P-Value | **RR (Confidence Interval)** |
| **Demographics** | Age Categories | 20-39 years | Reference | 1 |
| | | 40-49 years | 0.048 | 1.152 (1.002-1.325) |
| | | 50-69 years | 0.015 | 1.081 (1.016-1.150) |
| | | 70-100 years | 0.312 | 1.041 (0.963-1.126) |
| | Gender | Female | Reference | 1 |
| | | Male | 0.007 | 1.085 (1.022-1.151) |
| **Medical comorbidities** | COPD | | 0.111 | 0.955 (0.902-1.010) |
| | CHF | Absent | Reference | 1 |
| | | Present | 0.018 | 0.947 (0.905-0.991) |
| | Diabetes | | 0.568 | 0.980 (0.916-1.049) |
| | Hypertension | | 0.095 | 1.062 (0.990-1.140) |
| | Non-COVID-19 respiratory viral illness | Absent | Reference | 1 |
| | | Present | 0.000 | 0.908 (0.863-0.953) |
| | BMI | | 0.195 | 1.003 (0.998-1.008) |
| | Lymphocyte | | 0.771 | 1.009 (0.951-1.070) |
| | Asthma | | 0.411 | 1.048 (0.937-1.172) |
| | CKD | | 0.539 | 1.039 (0.920-1.173) |
| | SLE | | 0.167 | 0.945 (0.872-1.024) |
| | Organ Transplant | | 0.063 | 0.894 (0.793-1.006) |
| | CAD | | 0.711 | 1.015 (0.937-1.101) |
| | HIV | Absent | Reference | 1 |
| | | Present | 0.001 | 0.876 (0.813-0.944) |
| | Parkinson's disease | | 0.080 | 0.911 (0.822-1.011) |
| | Multiple sclerosis | | 0.148 | 0.949 (0.884-1.019) |
| | ESRD | Absent | Reference | 1 |
| | | Present | 0.012 | 0.909 (0.843-0.979) |
| | Stroke | | 0.383 | 0.968 (0.899-1.042) |
| | Demyelinating disease | | 0.297 | 0.932 (0.817-1.064) |
| | _cons | | 0.145 | 1.343 (0.904-1.997) |

Table T3 below shows the association between COVID-19 and age among never smokers. Male never smokers were more likely to test positive for COVID-19, while CHF, organ transplant, HIV positive, ESRD, non-COVID-19 respiratory viral illness patients were less likely to test positive for COVID-19.

| **Table T3: The Association between COVID-19 Status and Age among the never smokers** | | | | |
|---|---|---|---|---|
| | COVID-19 | | P-Value | RR (95% Conf. Interval) |
| **Demographics** | Age Categories | 20-39 years | Reference | 1 |
| | | 40-49 years | 0.212 | 1.101 (0.946-1.281) |
| | | 50-69 years | 0.045 | 1.078 (1.002-1.160) |
| | | 70-100 years | 0.829 | 1.009 (0.927-1.098) |
| | Gender | Female | Reference | 1 |
| | | Male | 0.008 | 1.010 (1.025-1.180) |
| **Medical comorbidities** | COPD | | 0.149 | 0.955 (0.896-1.017) |
| | CHF | Absent | Reference | 1 |
| | | Present | 0.007 | 0.931 (0.884-0.981) |
| | Diabetes | | 0.843 | 0.993 (0.922-1.068) |
| | Hypertension | | 0.084 | 1.074 (0.991-1.163) |
| | Non-COVID-19 respiratory viral illness | Absent | Reference | 1 |
| | | Present | 0.000 | 0.896 ( 0.847-0.949) |
| | BMI | | 0.704 | 1.001 (0.996-1.006) |
| | Lymphocyte | | 0.646 | 1.017 (0.945-1.096) |
| | Asthma | | 0.261 | 1.083 (0.943-1.243) |
| | CKD | | 0.529 | 1.042 (0.917-1.184) |
| | SLE | | 0.092 | 0.938 (0.870-1.011) |
| | Organ Transplant | Absent | Reference | 1 |
| | | Present | 0.024 | 0.860 (0.755-0.980) |
| | CAD | | 0.533 | 1.030 (0.937-1.134) |
| | HIV | Absent | Reference | 1 |
| | | Present | 0.001 | 0.875 (0.809-0.947) |
| | Parkinson's disease | | 0.171 | 0.917 (0.811-1.038) |
| | Multiple sclerosis | | 0.735 | 0.981 (0.879-1.095) |
| | ESRD | Absent | Reference | 1 |
| | | Present | 0.007 | 0.888 (0.814-0.969) |
| | Stroke | | 0.232 | 0.946 (0.865-1.036) |
| | Demyelinating disease | | 0.757 | 1.018 (0.908-1.142) |
| | _cons | | 0.308 | 1.243 (0.819-1.886) |

Table T4 below shows the association between COVID-19 and age among the current smokers. The 40 to 49 year old current smokers were more likely to test positive for COVID-19, while HIV positive, Non-COVID-19 respiratory viral illness current smokers while less likely to test positive for COVID-19.

| **Table T4: The Association between COVID-19 Status and Age among the current smokers** | | | | |
|---|---|---|---|---|
| | COVID-19 | | P-Value | RR [95% Conf. Interval] |
| **Demographics** | Age | 20-39 years | Reference | 1 |
| | | 40-49 years | 0.018 | 1.309 (1.047-1.635) |
| | | 50-69 years | 0.143 | 1.063 (0.980-1.153) |
| | | 70-100 years | 0.817 | 0.990 (0.907-1.080) |
| | Gender | | 0.053 | 1.064 (0.999-1.132) |
| **Medical comorbidities** | COPD | | 0.707 | 0.989 (0.933-1.048) |
| | CHF | | 0.061 | 0.943 (0.887-1.003) |
| | Diabetes | | 0.150 | 0.945 (0.874-1.021) |
| | Hypertension | | 0.104 | 1.080 (0.984-1.186) |
| | Non-COVID-19 respiratory viral illness | Absent | Reference | 1 |
| | | Present | 0.005 | 0.907 (0.846 -0.972) |
| | BMI4 | | 0.359 | 1.003 (0.997-1.008) |
| | Lymphocyt1e | | 0.270 | 1.035 (0.974-1.100) |
| | Asthma | | 0.758 | 1.021 (0.894-1.167) |
| | CKD | | 0.122 | 1.125 (0.969-1.308) |
| | SLE | | 0.188 | 0.918 (0.808-1.043) |
| | Organ Transplant | | | 1 |
| | CAD | | 0.592 | 1.023 (0.941-1.113) |
| | HIV | Absent | | |
| | | Present | 0.006 | 0.863 (0.775-0.959) |
| | Parkinson's disease | | 0.555 | 1.032 (0.930-1.144) |
| | Multiple sclerosis | | 0.098 | 0.898 (0.791-1.020) |
| | ESRD | | 0.092 | 0.936 (0.867-1.011) |
| | Stroke | | 0.680 | 0.980 (0.892-1.077) |
| | Demyelinating disease | | 0.147 | 0.929 (0.841-1.026) |
| | _cons | | 0.906 | 1.025 (0.678-1.549) |

Table T5 below shows the homogeneity test. The difference between never smokers and current smokers among the COPD, CHF, non-COVID-19 respiratory viral illness and Parkinson's disease is significantly different as shown by the homogeneity test in Supplementary table S3.

| **Table T5: The homogeneity test** | | | | |
|---|---|---|---|---|
| | COVID-19 | | P-Value | RR [95% Conf. Interval] |
| **Smoking Status** | Never-Current Smokers | | 0.434 | 0.973 (0.908- 1.042) |
| **Demographics** | Age Categories | 20-39 | Reference | 1 |
| | | 40-49 | 0.146 | 1.267 (0.921- 1.742) |
| | | 50-59 | 0.046 | 1.063 (1.001- 1.128) |
| | | 70-100 | 0.047 | 1.120 (1.001- 1.252) |
| | 2.NevCurSmokers | | Reference | 1 |
| | AgeCat6#NevCurSmokers | 20 2 | Reference | 1 |
| | | 40 2 | 0.142 | 0.782 (0.563- 1.086) |
| | | 50 2 | 0.837 | 1.011 (0.911- 1.122) |
| | | 70 2 | 0.728 | 0.975 (0.844- 1.126) |
| | Gender#c.NevCurSmokers | | Reference | 1 |
| | | Male | 0.096 | 1.044 (0.992- 1.098) |
| **Medical comorbidities** | COPD#c.NevCurSmokers | | 1 | Reference |
| | | Yes | 0.006 | 0.921 (0.868- 0.977) |
| | CHF#c.NevCurSmokers | | 1 | Reference |
| | | Yes | 0.029 | 0.959 (0.923- 0.996) |
| | Diabetes | | 0.602 | 1.023 (0.940- 1.112) |
| | Hypertension | | 0.541 | 1.026 (0.945- 1.113) |
| | Viral1#c.NevCurSmokers | | Reference | 1 |
| | | 2 Yes | 0.003 | 0.950 (0.918 - 0.983) |
| | BMI4 | | 0.086 | 1.006 (0.999- 1.013) |
| | Lymphocyt1e | | 0.988 | 1.001 (0.925- 1.082) |
| | Asthma | | 0.553 | 1.038 (0.918- 1.172) |
| | CKD | | 0.058 | 0.907 (0.820- 1.003) |
| | SLE | | 0.247 | 0.817 (0.580- 1.150) |
| | Organ Transplant | | 0.262 | 0.930 (0.820- 1.056) |
| | CAD | | 0.891 | 0.993 (0.902- 1.094) |
| | HIV | | Reference | 1 |
| | Parkinson's disease | | 0.006 | 0.819 (0.709- 0.945) |
| | Multiple sclerosis | | 0.605 | 1.034 (0.911- 1.174) |
| | ESRD#c.NevCurSmokers | | Reference | 1 |
| | | Yes | 0.311 | 0.966 (0.904- 1.033) |
| | Stroke | | 0.110 | 0.945 (0.882- 1.013) |
| | Demyelinating disease | | 0.153 | 0.923 (0.826- 1.030) |

### Abbreviations

ACE-2 angiotensin converting enzyme 2
CAD coronary artery disease
CDC center for disease control
COPD chronic obstructive pulmonary disease
BMI body mass index
SLE systemic lupus erythromatosus
CHF congestive heart failure
ESRD end stage renal disease
CKD chronic kidney disease
COVID-19 corona virus disease 2019
HIV human immunodeficiency virus
Interquartile range IQR
Polymerase chain reaction PCR
RSV respiratory syncytial virus
USA united states of America
UK United Kingdom
